# Bail-out transvenous temporary pacing during rotational atherectomy PCI

**DOI:** 10.1101/2023.10.12.23296980

**Authors:** Konstantin Schwarz, Julia Mascherbauer, Elisabeth Schmidt, Martina Zirkler, Paul Vock, Gudrun Lamm, Chun Shing Kwok, Josip Andelo Borovac, Roya Anahita Mousavi, Uta C. Hoppe, Gregor Leibundgut, Maximilian Will

**Author notes:** **Corresponding author** Dr. Konstantin Schwarz, University Hospital St. Pölten, Dunant-Platz 1, 3100 St. Pölten, Austria.

## Abstract

**Background:** Rotational atherectomy (RA) percutaneous coronary intervention (PCI) may cause transient bradycardia or heart block. Traditionally, some operators use prophylactic transvenous pacing wire (TPW) to avoid haemodynamic complications associated with bradycardia. We sought to establish the frequency of bail-out need for emergency TPW insertion in patients undergoing RA PCI that have received no upfront TPW insertion.

**Methods:** We performed a single-centre retrospective study of all patients undergoing RA PCI between October 2009 and October 2022. Patient characteristics, procedural variables and in-hospital complications were registered.

**Results:** A total of 331 patients who underwent RA procedure were analyzed. No patients underwent prophylactic TPW insertion. The mean age was 73.3±9.1 years, 71.6% (n=237) were male, while nearly half of patients were diabetic (N=47.7%, N=158). The right coronary artery was the most common target for RA PCI (40.8%), followed by left anterior descending (34.1%), left circumflex (14.8%) and left main stem artery (10.3%). Twenty (6%) of patients required intraprocedural atropine therapy. Emergency TPW insertion was needed in one patient (0.3%) only. Eight (2.4%) patients died, however only one was adjudicated as possibly related to RA-induced bradycardia. Five patients (1.5%) had ventricular fibrillation arrest while nine (2.7%) required cardiopulmonary resuscitation. Six (1.8%) procedures were complicated by coronary perforation, two (0.6%) were complicated by tamponade while 17 (5.1%) patients experienced vascular access complications.

**Conclusions:** Bail-out transvenous temporary pacing is very rarely required during RA PCI. A standby temporary pacing strategy is reasonable and may avoid unnecessary TPW complications compared to routine use.

## INTRODUCTION

Rotational atherectomy (RA) plays a pivotal role in contemporary calcium modification armamentarium during percutaneous coronary intervention (PCI). This technique was first described by Fourrier in a series of 12 patients in 1989 (1). Despite technical improvements over time in equipment and technology, the basic characteristics of the procedure have remained largely unchanged. However, with increasing experience among operators, various recommendations of the practical handling of the device have evolved. Compared to bail-out RA PCI strategy, upfront planned RA decreases peri-procedural complications (2).

Further, it was noticed that due to risk of distal embolization of calcific microparticles and associated neuro-hormonal response, a transient heart block can occur. This reaction was noticed more frequently following RA of right coronary artery or dominant circumflex artery (3). Lesion length, burr-to-artery ratio and total duration of runs were other independent predictors of RA induced bradycardia (4).

Historically, a routine prophylactic transvenous temporary pacing wire (TPW) was recommended for prevention of symptomatic bradycardia induced by rotational atherectomy. Mitar et al. described in a small single centre retrospective observational study (n=134) use of TPW in 50% of patients and in 42 (31%) patients heart block or pacemaker activation was described (3).

Over the last decade, clinical practice turned away from prophylactic TPW insertion and many experts now recommend no routine prophylactic TPW insertion prior to procedures involving RA (5, 6). Currently, there is a large paucity of data supporting or disproving this strategy. Concerns of temporary pacing wire complications on one hand, and the relative short-lived bradycardias and AV blocks (which respond quickly to vagolytic manoeuvres) on the other hand may favour the conservative strategy.

In this single-centre study, we sought to establish the real-life occurrence of significant bradycardia and AV block necessitating temporary pacing wire insertion during RA PCI. RA PCI is usually performed in coronary lesions with severe calcifications, complex anatomy, frail high-risk patients and carries higher risk of death, cardiac tamponade or emergency bypass surgery (7). Predictors of RA PCI complications are patient related (old age, impaired kidney function or previous myocardial infarction) and procedure related (emergency procedure, triple-vessel disease and low institutional volume) (7). As a secondary outcome we elected to document other procedural complications related to high-risk PCI including coronary perforation, tamponade, stalled RA burr, cardiac arrest, emergency CABG, death and vascular access complications.

## METHODS

This retrospective observational single-centre study was performed at a tertiary hospital centre, the University Hospital Sankt Pölten, Austria. It was approved the Karl-Landsteiner scientific integrity and ethics commission (ethics commission number 1057/2023) and was performed in compliance with the Declaration of Helsinki. The reporting of this study is in accordance to the STrengthening the Reporting of OBservational studies in Epidemiology (STROBE) recommendations (8).

All patients undergoing RA PCI between 1^st^ of October 2009 and 31^st^ of October 2022 in our centre were identified from our hospital PCI database. The primary aim of this study was a description of the frequency of bradycardia and AV block necessitating bail-out TPW insertion during RA procedure at our centre.

### Statistical analysis

All analyses were performed using MS Excel 2016 (Microsoft, Redmond, CA) and GraphPad Prism (Version 5.04) software. Descriptive statistics were performed. Categorial variables were expressed as absolute numbers and percentages. Continuous variables were expressed as mean and standard deviation or median and interquartile range, depending on the normality of distribution. If a larger number of emergency TPW procedures were to be identified, then an exploratory analysis examining possible predictors for TPW was planned.

## RESULTS

In total, 331 patients undergoing RA PCI were identified from our hospital database. Prophylactic temporary pacing wire insertion prior to planned RA in a stable patient has not been routine practice in our centre. Hence no patient in our sample received temporary wire as primary prevention of symptomatic bradycardia prior to RA.

Two-hundred and thirty-seven patients (71.6%) were male and the majority of procedures were performed in elective setting (N=207, 60.7%). Twenty-two (6.6%) patients had permanent pacing system implanted before RA. Right coronary artery (RCA) (40.8%) and left anterior descending (LAD) (34.1%) were the most commonly treated RA target vessels. Femoral access was used in 161 (48%) patients. Detailed demographics of patient population are shown in **Table 1**. Intraprocedural complications are shown in **Table 2**.

**TABLE 1.** Demographics and procedural characteristics of patients undergoing rotational atherectomy. SD standard deviation, IQR interquartile range, BMI body mass index, LV left ventricle, PM pacemaker, ICD implantable cardioverter defibrillator, CRT cardiac resynchronisation therapy, CKD chronic kidney disease, PAD peripheral artery disease, CABG coronary artery bypass grafting, CTO chronic total occlusion, NSTEMI non-ST elevation myocardial infarction, UA unstable angina

| **Demographics and procedural characteristics**<br>(n=331) | **n (%) or mean±SD or median [IQR]** |
| --- | --- |
| Age | 73.3 ± 9.1 |
| Male sex, n (%) | 237 (71.6) |
| Height, cm (SD) | 170.8 ± 6.5 |
| Weight, kg (SD) | 89.2 ± 9.8 |
| BMI, mean (SD) | 29.9 (2.0) |
| History of congestive heart failure, n (%) | 125 (37.8) |
| LV function, n (%) |  |
| normal | 207 (60.7) |
| Mildly impaired | 56 (16.9) |
| Moderately impaired | 40 (12.1) |
| Severely impaired | 28 (8.5) |
| Valvular pathology | 100 (30.2) |
| Permanent PM/ICD/CRT | 22 (6.6) |
| Diabetes n (%) | 158 (47.7) |
| CKD | 70 (21.1) |
| Creatinine, median (IQR) | 0.97 [0.83-1.25] |
| Stroke/TIA | 69 (20.8) |
| Hypertension | 297 (89.7) |
| Smoker | 159 (48.0) |
| current | 62 (18.7) |
| Ex-smoker | 97 (29.3) |
| Dyslipidaemia | 280 (84.6) |
| PAD | 66 (19.9) |
| CABG history | 64 (19.3) |
| Multivessel disease | 305 (92.1) |
| CTO | 18 (5.4) |
| Presentation |  |
| Elective | 205 (61.9) |
| NSTEMI/UA | 87 (26.3) |
| STEMI | 39 (11.8) |
| Access |  |
| radial | 163 (49.2) |
| femoral | 161 (48.6) |
| brachial | 7 (2.1) |
| Artery treated with RA |  |
| RCA | 135 (40.8) |
| LAD | 113 (34.1) |
| CX | 49 (14.8) |
| LM | 34 (10.3) |

**TABLE 2.**
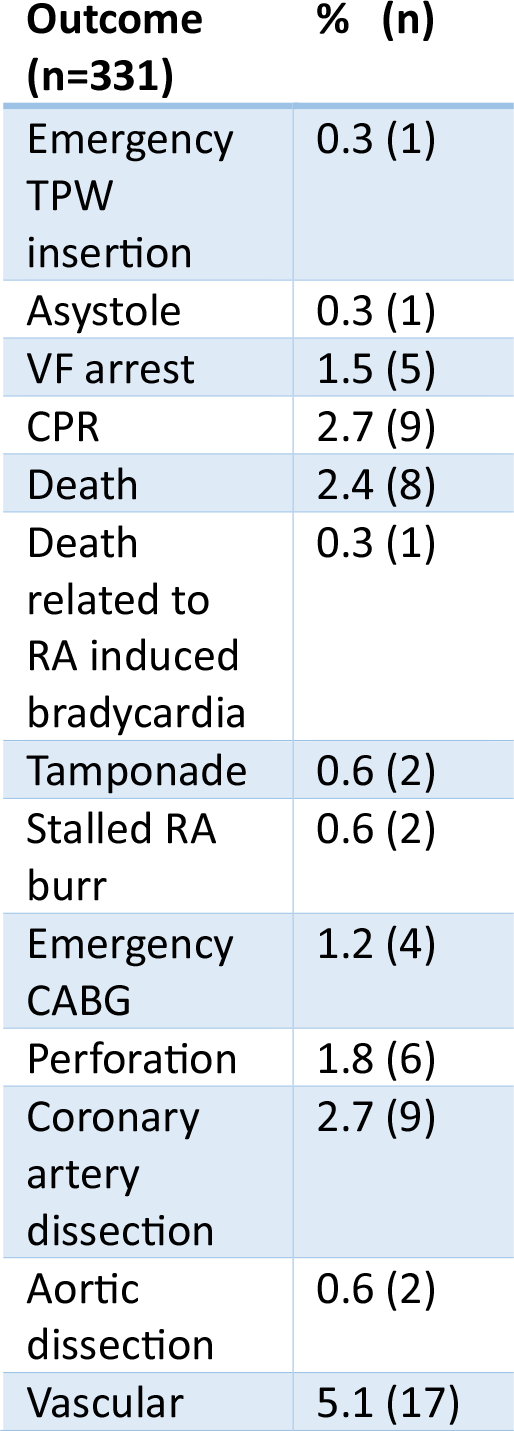
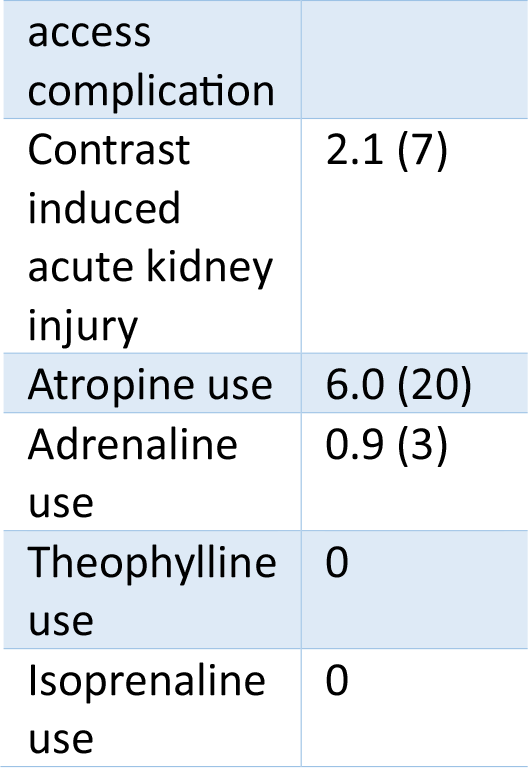
Outcomes in patients undergoing rotational atherectomy. TPW transvenous pacing wire, VF ventricular fibrillation, CPR cardiopulmonary resuscitation, RA rotational atherectomy, CABG coronary artery bypass graft

Asystole was documented in 1 (0.3%) patient. Administration of intravenous atropine during procedure occurred in 20 (6%) patients. Three (0.9%) patients received adrenaline. Only one (0.3%) patient received an emergency temporary pacing wire. Nine (2.7%) patients have required cardiopulmonary resuscitation, 5 (1.5%) had VF arrest while 8 (2.4%) patients died following RA PCI. However, it was only one (0.3%) patient (the same who received the emergency temporary pacing wire) whose death is likely to be directly contributed to by RA-induced bradycardia. The patient that died underwent rotational atherectomy due to a severely calcified LAD, in the concomitant presence of RCA CTO.

Other complications, such as stalled burr, coronary perforation, emergency coronary artery bypass graft (CABG) or aortic dissection were cumulatively relatively rare (<3% in total). Vascular access complications were reported in 17 (5.1%) of cases. Due to the low number of the primary outcome (bail-out TPW insertion; n=1), no further statistical tests to identify predictors of TPW insertion were performed.

## DISCUSSION

Rotational atherectomy is an important PCI technique utilized for calcium modification in complex coronary artery disease. Its use varies from <1% up to 10% of all PCIs in selected centres (5). In Japan (6) or United Kingdom it is used in about 3 % of all PCI procedures, whereas in Germany in only approximately 0.8% of all PCI cases (9). In United Kingdom, the number of RA procedures is continuously declining since 2018, which is likely related to the introduction and continuous increase of intravascular lithotripsy use over the last few years (10).

Our study shows that RA is performed in high-risk population with almost half of all patients (47.7%) being diabetic and/or having history of smoking, obesity with significant proportion of patients having chronic kidney disease (CKD) (21%), peripheral artery disease (PAD) (20%) and advanced coronary artery disease (92% of cases were having multi-vessel disease). More than one third of all patients presented with acute coronary syndrome (ACS) or ST-elevation myocardial infarction (STEMI). These characteristics are very similar to a ROTATE multicentre registry published by Kawamoto et. Al (11). Females patients were previously shown to be older, have more frequently femoral access and experience more net adverse clinical events compared to male patients undergoing RA PCI (12).

In our centre, the rate of radial access for most operators ranges between 85 to 90%. There was a high proportion of femoral access (48%) in the RA PCI group which was likely caused by two factors. First, all procedures performed before 2013 were performed by femoral access, reflecting the historical practice in Austria at that time. This theory would be supported by a sub-analysis of our center’s RA PCI procedures since the year 2020, when the rate of femoral access use decreased to 27% and the vast majority of procedures were performed via trans-radial route. A multi-centre study from United Kingdom enrolling 518 patients treated with RA between 2005 and 2013 confirmed a similar notion when at the time only 30.7% of all RA procedures were performed *via* trans-radial approach (13).

Secondly, as mentioned above, the population which undergoes RA PCI is per definition a high-risk population with substantial prevalence of PAD and CKD, which is likely affecting the peripheral access options. The nature of heavily diseased coronary arteries, better support and the need to accommodate bigger RA burrs frequently necessitates larger guides (*e*.*g*. 7 French) and hence femoral access, compared to non-RA PCI.

The main finding of our study is that despite the occasionally documented transient bradycardias, which necessitated the use of pharmacological treatment (atropine was used in 6% of patients), the need for emergency TPW insertion was exceedingly rare. Only one patient (0.3%) required emergency temporary wire. Unfortunately, this patient suffered cardiac arrest and died, and the RA induced bradycardia may have played an important role in this death. In-hospital mortality occurred in 2.7% (n=8) of patients. The vast majority of the deaths (except one) were not related to bradycardia induced by the RA procedure. Likewise, all other intraprocedural complications were very rare as previously shown by our data.

Periprocedural and in-hospital mortality in our study, compares unfavourably with mortality seen in elective uncomplicated PCI cases which is usually below 1% (BCIS UK 0.2%), however it is in line with deaths reported for all PCI (2.3 %) or primary PCI (6.2%) [PCI mortality data in brackets taken from BCIS audit 2021/2022 (10)]. Eftychiu et al. reported, in a population of patients treated with RA that presence of PAD, DM, ACS presentation and SYNTAX Score ≥23 were all independent predictors of major adverse events (13). Pharmacological bail-out strategies were occasionally needed in our population (atropine in 6% and Adrenaline in 0.3%) to treat significant bradycardias. Theophyline or aminophylline was not used in our institution, however is occasionally used in other institutions to treat or prevent RA PCI related bradycardia. Acar et al demonstrated in a small open label study (n=60) a reduction of bradycardia and heart block with a prophylactic aminophylline added to heparin and nitroglycering saline flush solution (rotaphilline) when compared to heparin and nitroglicerin only saline flush solution (4).

Our finding of the exceedingly rare requirement for bail out emergency temporary pacing implantation (0.3%), in an unselected real-life population undergoing PCI, is a central theme of our study. It supports the clinical experience of operators over the last decade and supports the increasingly prevalent expert consensus (currently not based on any data), that routine temporary wire insertion prior to RA is not necessary (5, 9).

The rare need for TPW appears in contrast to the findings of Mitar et al (3), where 50% of patients had TPW inserted as primary prevention measure. The authors reported that 31% of patients developed higher AV block or experienced TPW activation during the RA procedure. Whereas a transient AV block or bradycardia is fairly common during RA PCI, in our experience, usually non-invasive vagolytic manoeuvres are sufficient to treat a vast majority, if not all, of these situations. In our single-centre study, there was only one case of emergency TPW insertion, which unfortunately led to death and it is possible this death was related to the bradycardia induced by the RA. However, the risk-benefit ratio has to be weighed against potential risks of 330 other procedures where TPW insertion was avoided and hence potentially prevented TPW-associated risks that are potentially lethal (*e*.*g*. RV wall perforation, cardiac tamponade, vascular access complications, and death). In other words, the theoretical number-needed-to-treat (NNT) appears to be very high for the prophylactic TPW. A recent review by Tjong et al. reported that in the period from 2010 to 2019, transvenous pacing for a variety of clinical scenarios was associated with a mean complication rate of 22.9%, out of which 5.7% were considered clinically significant (14). Another consideration is the additional procedural time and the material cost associated with routine prophylactic TPW insertion.

There are several techniques which can minimize the risk of significant bradycardias, or provide alternative, less invasive strategies compared to temporary pacing wire insertion. These include start of the procedure with smaller burrs, use of lower rotational speed, sufficient pauses between individual runs, vagolytic manouvers (cough), pharmacology (atropine, adrenaline, aminophylline) and safety setups including routine external pacing pads and washed femoral access for emergency bail out temporary wire insertion if needed (4, 15). An important role to minimize major adverse events associated with RA plays the experience of the operator. Kinnaird et al. showed that in-hospital mortality and adverse events occurred less frequently as operator RA PCI volume increased (16).

Another interesting alternative to TPW pacing is the trans-coronary pacing (TCP) over the rota wire which was first described by Meier in 1985 (17, 18). Recently, Iqbal et all reported results of prophylactic trans-coronary pacing in ROTA-PACE study in 132 patients. Successful TCP was reported in 121 (91.7%) patients and no complications were registered (19). The ROTA-PACE technique was not successful in 11 patients, out of which in four the rota wire could not be placed distally and four patients had large infarcts in the territory of the rota wire. A further development of the trans-coronary pacing is the use of Electroducer sleeve developed recently by Benjamin Faurie. This is a special electro-conductive sheath which is supposed to standardise the direct wire pacing technique (both for TAVI or PCI) with more reproducibility and acceptable thresholds (20).

Future prospective randomized studies are warranted to investigate the safety and efficacy of systematic use of prophylactic pharmacology together with bail-out trans-coronary pacing compared vs. bail-out transvenous pacing vs. routine prophylactic transvenous pacing in patients undergoing RA PCI.

### Limitations

There are several limitations to this study. The main limitation is the retrospective single-centre nature of this study and the fairly long observational period included. There is likely a wide variety of experiences of different operators with the RA procedure. This may have played an important role especially in the first few years of the observational period. However, it is extremely reassuring, that the bail out need for routine transvenous pacing was exceedingly rare.

## Conclusion

We present that the bail out transvenous temporary pacing is rarely needed during RA PCI. A standby temporary pacing strategy is reasonable and may avoid unnecessary procedural complications when compared to routine prophylactic transvenous pacing strategy. Multiple non-invasive strategies and optimisation of the RA technique may minimize the risk of clinically significant bradycardias during RA PCI. An individualized approach with TPW wire insertion or the use of trans-coronary pacing via the rota wire in very high-risk Patients may offer reasonable alternatives to routine transvenous temporary pacing wire insertion.

## Data Availability

No data from publically available sources were used.

## Acknowledgments

The authors want to appreciate the contribution of NÖ Landesgesundheitsagentur, legal entity of University Hospitals in Lower Austria, for providing the organizational framework to conduct this research. The authors also would like to acknowledge support by Open Access Publishing Fund of Karl Landsteiner University of Health Sciences, Krems, Austria.

---

**Figure 1.**
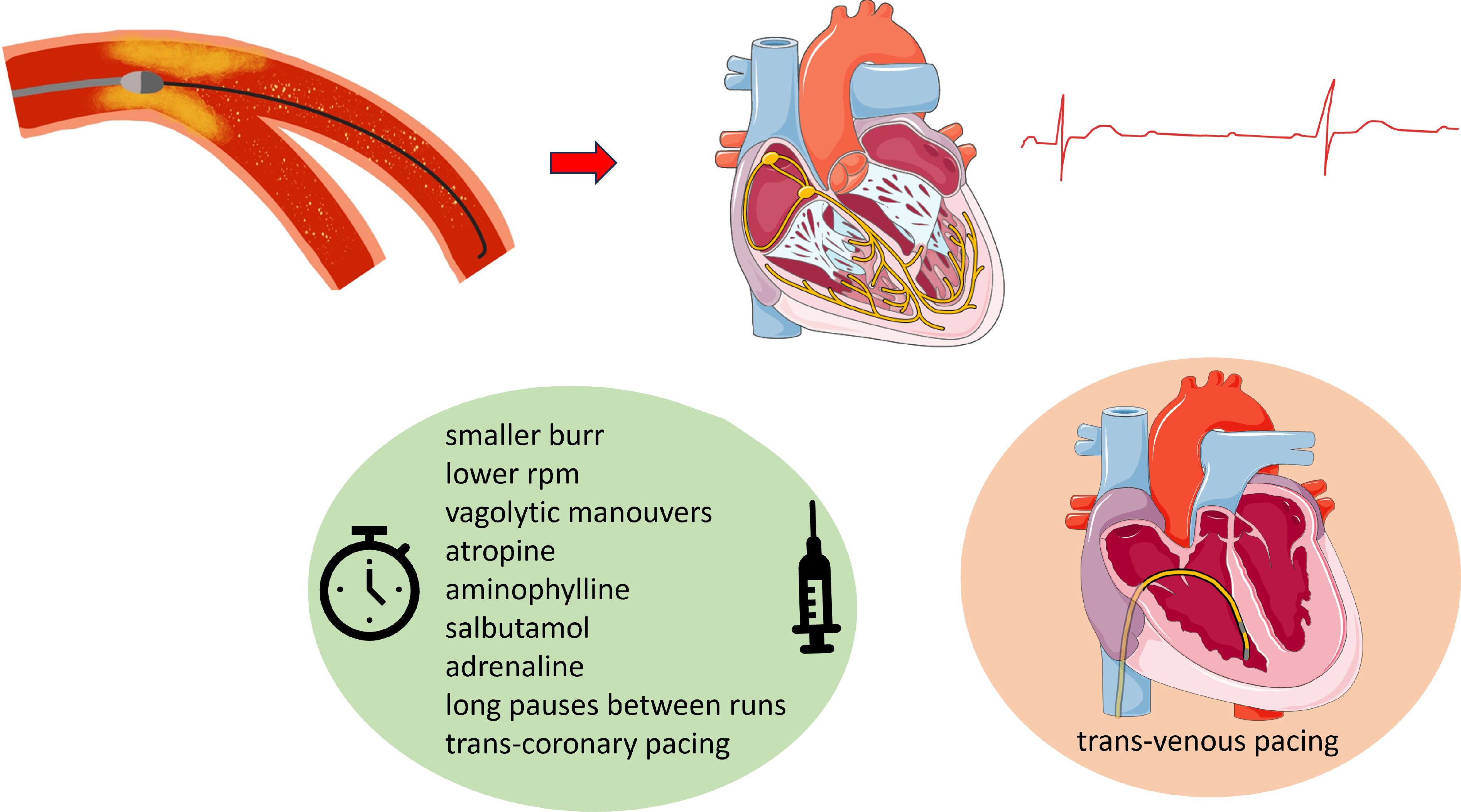
Rotational atherectomy induced microembolisation can induce reflective bradycardia and transient heart block. There are several procedural aspects, which can limit the occurrence or impact of symptomatic bradycardia. *Figures were partly adapted and by courtesy from Servier Medical Art*.

## Abbreviations used

PCI: percutaneous coronary intervention
RA: rotational atherectomy
TPW: temporary pacing wire
RCA: right coronary artery
CX: circumflex coronary artery
LAD: left anterior descending artery
LM: left main stem
DM: diabetes mellitus
PAD: peripheral artery disease
CKD: chronic kidney disease
CPR: cardiopulmonary resuscitation
VF: ventricular fibrillation

## REFERENES

1. Fourrier JL, Bertrand ME, Auth DC, Lablanche JM, Gommeaux A, Brunetaud JM. Percutaneous coronary rotational angioplasty in humans: Preliminary report. J Am Coll Cardiol. 1989;14(5):1278–82.

2. Schwarz K, Lovatt S, Borovac JA, Parasuraman S, Kwok CS. Planned Versus Bailout Rotational Atherectomy: A Systematic Review and Meta-Analysis. Cardiovasc Revasc Med. 2022;39:45–51.

3. Mitar MD, Ratner S, Lavi S. Heart block and temporary pacing during rotational atherectomy. Can J Cardiol. 2015;31(3):335–40.

4. Acar E, Izci S, Donmez I, Ozgul N, Ozcan E, Kaygusuz T, et al. A mix of aminophylline and heparin plus nitroglycerin can reduce bradycardia during rotational atherectomy on the right coronary artery and dominant circumflex artery. Herz. 2023.

5. Sharma SK, Tomey MI, Teirstein PS, Kini AS, Reitman AB, Lee AC, et al. North American Expert Review of Rotational Atherectomy. Circ Cardiovasc Interv. 2019;12(5):e007448.

6. Sakakura K, Ito Y, Shibata Y, Okamura A, Kashima Y, Nakamura S, et al. Clinical expert consensus document on rotational atherectomy from the Japanese association of cardiovascular intervention and therapeutics: update 2023. Cardiovascular Intervention and Therapeutics. 2023;38(2):141–62.

7. Sakakura K, Inohara T, Kohsaka S, Amano T, Uemura S, Ishii H, et al. Incidence and Determinants of Complications in Rotational Atherectomy: Insights From the National Clinical Data (J-PCI Registry). Circ Cardiovasc Interv. 2016;9(11):e004278.[pii].

8. von Elm E, Altman DG, Egger M, Pocock SJ, Gøtzsche PC, Vandenbroucke JP. The Strengthening the Reporting of Observational Studies in Epidemiology (STROBE) statement: guidelines for reporting observational studies. Lancet. 2007;370(9596):1453–7.

9. Barbato E, Carrié D, Dardas P, Fajadet J, Gaul G, Haude M, et al. European expert consensus on rotational atherectomy. EuroIntervention : journal of EuroPCR in collaboration with the Working Group on Interventional Cardiology of the European Society of Cardiology. 2015;11(1):30–6.

10. Society BCI. BCIS Audit 2021-2022. 2023.

11. Kawamoto H, Latib A, Ruparelia N, Boccuzzi GG, Pennacchi M, Sardella G, et al. Planned versus provisional rotational atherectomy for severe calcified coronary lesions: Insights From the ROTATE multi-center registry. Catheter Cardiovasc Interv. 2016;88(6):881–9.

12. Ford TJ, Khan A, Docherty KF, Jackson A, Morrow A, Sidik N, et al. Sex differences in procedural and clinical outcomes following rotational atherectomy. Catheter Cardiovasc Interv. 2020;95(2):232–41.

13. Eftychiou C, Barmby DS, Wilson SJ, Ubaid S, Markwick AJ, Makri L, et al. Cardiovascular Outcomes Following Rotational Atherectomy: A UK Multicentre Experience. Catheter Cardiovasc Interv. 2016;88(4):546–53.

14. Tjong FVY, de Ruijter UW, Beurskens NEG, Knops RE. A comprehensive scoping review on transvenous temporary pacing therapy. Neth Heart J. 2019;27(10):462–73.

15. Safian RD, Feldman T, Muller DW, Mason D, Schreiber T, Haik B, et al. Coronary angioplasty and Rotablator atherectomy trial (CARAT): immediate and late results of a prospective multicenter randomized trial. Catheter Cardiovasc Interv. 2001;53(2):213–20.

16. Kinnaird T, Gallagher S, Sharp A, Protty M, Salim T, Ludman P, et al. Operator Volumes and In-Hospital Outcomes: An Analysis of 7,740 Rotational Atherectomy Procedures From the BCIS National Database. JACC Cardiovasc Interv. 2021;14(13):1423–30.

17. Meier B, Rutishauser W. Coronary pacing during percutaneous transluminal coronary angioplasty. Circulation. 1985;71(3):557–61.

18. Kusumoto H, Ishibuchi K, Hasegawa K, Otsuji S. Trans-coronary pacing via Rota wire prevents bradycardia during rotational atherectomy: a case report. Eur Heart J Case Rep. 2022;6(2):ytac013.

19. Iqbal MB, Robinson SD, Nadra IJ, Das D, van Zyl M, Sikkel MB, et al. The Efficacy and Safety of an Adjunctive Transcoronary Pacing Strategy During Rotational Atherectomy: ROTA-PACE Study. JACC Cardiovasc Interv. 2023.

20. Wintzer-Wehekind J, Lefèvre T, Benamer H, Monsegu J, Tchétché D, Garot P, et al. A direct wire pacing device for transcatheter heart valve and coronary interventions: a first-in-human, multicentre study of the Electroducer Sleeve. EuroIntervention : journal of EuroPCR in collaboration with the Working Group on Interventional Cardiology of the European Society of Cardiology. 2023;18(14):1150–555.

